# A new indicator to measure discordance between patient reported outcomes and traditional disease activity holds promise to advance care trajectories improve care in patients with early Rheumatoid Arthritis

**DOI:** 10.1101/2021.12.01.21267120

**Authors:** Sofia Pazmino, Anikó Lovik, Annelies Boonen, Diederik De Cock, Veerle Stouten, Johan Joly, Michaël Doumen, Delphine Bertrand, René Westhovens, Patrick Verschueren

## Abstract

**Objective:** To unravel disease impact in early RA patients by separately quantifying patient reported (PRF), clinical (CF) and laboratory (LF) factors. We put forward a new indicator, the discordance score (DS), for early identification and prediction of unmet patient outcomes in terms of future achievement of sustained remission and RA-related quality of life (QoL).

**Methods:** We obtained factor scores via factor analysis in the CareRA trial, then calculated the DS between PRF and the mean of the other scores. We computed the improvement from baseline to week 104 (%) and area-under-the-curve (AUC) across time-points per factor score and compared these between patients achieving or not achieving sustained (week 16 to 104) remission (DAS28CRP<2.6) with ANOVA. Logistic and linear regressions respectively were used to predict SR based on previous factor and discordance scores, and QoL at year 1 and 2 based on DS at week 16.

**Results:** PRF, CF and LF scores improved rapidly within 8 weeks. In patients achieving SR; PRF improved 57%, CF 90% and LF 27%, compared to 32% PRF (p=0.13), 77% CF (p<0.001) and 9% LF (p=0.36) score improvement in patients not achieving SR. Patients achieving SR had an AUC of 15.7, 3.4 and 4.8 for PRF, CF and LF respectively, compared to 33.2, 10.1, and 7.2 in participants not achieving SR (p<0.001 for all). Early factor and discordance scores were associated with later stage factor scores as well as QoL and PRF scores predicted SR (p<0.005 for PRF and DS).

**Conclusions:** All factor scores improved rapidly, especially in patients achieving SR. Patient-reported burden improved less extensively. Discordance scores could help in predicting the need for additional non-pharmacological interventions to achieve SR and decrease disease impact.

**KEY MESSAGES:** *What is already known about this subject?:*

- Early and intensive RA drug-treatment using disease activity as a target allows rapid disease control and prevents joint destruction.
- Including pain, fatigue and physical function when assessing patients with early RA broadens the evaluation of disease impact.

*What does this study add?:*

- Leveraging patient reported outcomes (pain, fatigue and physical function) and traditional disease activity measures, we introduce a new indicator (named discordance score) for unraveling disease impact and treatment efficacy.
- We show how the discordance score stands for current unmet patient reported outcomes and could be used to predict future sustained disease contol and quality of life (1 and 2 years after baseline).
- We demonstrate this effect both in patients with and without sustained remission

*How might this impact on clinical practice or future developments?:*

- The earlier detection of unmet needs despite good disease control could allow to perform timely interdisciplinary interventions other than medication adaptations and could promote psychosocial wellbeing for patients.

## INTRODUCTION

The primary clinical manifestation of Rheumatoid Arthritis (RA) is inflammation of the peripheral joints resulting in swelling, stiffness and pain.^1^ However, more constitutional symptoms such as fatigue, pain and stiffness, as well as a restricted ability to work, and impact on other aspects of health-related quality of life (QoL) can be present.^2^ Treating early to a target of remission or at least low disease activity (LDA) is highly advocated.^3^ This treatment strategy aims to maximise the long-term health-related quality of life (HR-QoL) through control of symptoms, prevention of structural damage, normalisation of function and social participation.^4^ A clear definition of target is paramount and should be stringent enough to seek “the best possible control of inflammation” while avoiding overtreatment.^5^

Specific instruments are used to measure the target of remission or LDA.^3^ For example, the ACR/EULAR Boolean remission criterion is stringent, requiring swollen/tender joint counts (SJC/TJC) to be below or equal to (≤) 1, C-reactive protein (CRP) ≤1mg/dL and Patient’s global health (PaGH) ≤1 (0-10 scale). When using this criterion for remission, it has been shown that one-third of RA patients fail to reach remission solely because of PaGH (near-remission).^6^ If the current treatment recommendations would be followed,^3^ based on the Boolean remission definition a state of near-remission could lead to an adaptation of disease modifying anti-rheumatic drugs (DMARDs), although isolated PaGH elevation suggests needs that are not necessarily related to inflammation.^7^ Hence, in such cases the remaining disease burden might not, or not only be mediated by disease activity. Several studies have shown a statistically significant correlation of PaGH with disease activity;^7,8^ however, this correlation is absent in low levels of disease activity where PaGH is related to pain, function and fatigue.^7–9^ Unmet needs incorporated in the PaGH and their relative importance should be uncovered when aiming to reduce the broader impact of RA.

Recent research has shown that adding pain, fatigue and physical function to the components included in the traditional composite disease activity measures may provide valuable additional information in evaluating the disease impact on the patient while keeping the traditional components intact.^10^ Three factors were identified: a patient-reported (PRF), clinical (CF) and laboratory (LF) factor. PRF consists of PaGH, pain, fatigue and physical function, so four self-reported measures that add a separate patient-experienced aspect of evaluating the disease burden. CF contains three variables: the physician’s global health assessment (PhGH), swollen and tender joint counts, all of which are common clinical measures. LF is comprised by only two laboratory measures, CRP and ESR. CF and LF scores have been part of measuring disease activity for a long time, while PRF is a novel addition. The three factors have also been replicated in a patient sample with established RA.^11^ Based on these results this paper aims to explore the potential benefit of evaluating these factor scores and their patterns over time in the CareRA trial^12^ for better understanding of the disease burden from the patient’s perspective. For this reason, we will plot the evolution of the three factors and take their differences under the loop with a new indicator called <u>the discordance score</u> which allows us to study the discordance between the traditional disease activity measures and patient-reported outcomes. We will attempt to predict from the baseline PRF, CF, LF and discordance scores both later scores and QoL outcomes. Moreover, by comparing patients achieving sustained remission (SR) -from week 16 until the end of this 2-year trial- to those who do not achieve sustained remission, we illustrate how even the patients achieving SR still report unmet needs such as pain and fatigue.^13^

## PATIENTS AND METHODS

CareRA was a 2-year open-label investigator-initiated pragmatic superiority trial (EudraCT number: 2008-007225-39, Clinical trials NCT01172639) conducted in 13 Flemish rheumatology centres.

Patients with recently diagnosed RA (≤1 year) were included and stratified into a high- or low-risk group based on classical factors of poor prognosis (erosions, rheumatoid factor (RF) and/or anti-citrullinated cyclic peptide (anti-CCP) positivity) and baseline disease activity score in 28 joints with C-reactive protein (DAS28CRP) >3.2) and then randomised into four different treatment arms. High-risk patients were randomised to methotrexate (MTX) 15mg weekly with a step-down glucocorticoid (GC) scheme or to this combination together with either sulphasalazine or leflunomide. Low-risk patients were randomised to a tight step-up treatment starting with MTX monotherapy without GC or to MTX weekly with step-down GCs. Overall, around 70% of the CareRA participants achieved a status of good disease control after 2 years (DAS28CRP <2.6) with a treat-to-target approach.^12^

### Clinical outcomes

Patients were assessed at screening, baseline and further at week 8, 16, 28, 40, 52, 65, 78, 91 and 104. Optional visits, if clinically required, could be performed. An electronic case report form was filled out and routinely monitored. Clinical, patient and laboratory parameters were collected at every visit: swollen (SJC28) and tender joint (TJC28) count in 28 joints, patient’s global health assessment (PaGH), physician’s global health assessments (PhGH), C-reactive protein (CRP) or erythrocyte sedimentation rate (ESR), health assessment questionnaire (HAQ), pain and fatigue each on a visual analogue scale (VAS) of 0-100. Overall QoL and RA-specific QoL (RAQoL) was captured with the Short Form 36 (SF-36) and RAQoL questionnaire at baseline, week 16, 52 and 104. The SF-36 was standardized in 1990 as a self-report measure of functional health and well-being. The SF-36 consists of eight scales: physical functioning (10 items), role-physical (4 items), bodily pain (2 items), general health (5 items), vitality (4 items), social functioning (2 items), role-emotional (3 items), mental health (5 items), and a final item, termed self-reported health transition. To score the SF-36, scales are standardized with a scoring algorithm to obtain a score ranging from 0 to 100.^14^ Higher scores indicate better health status. The RAQoL contains 30 yes/no questions regarding specific activities of daily living and quality of life. Each positive response is one point, a total sum is calculated giving a scale of 0-30.^14^ Higher scores indicate worse health status.

### Statistical analyses

All randomised patients who had taken at least one medication dose, were considered for the intention to treat (ITT) analysis. Missing data were imputed with multiple imputation. As described previously by Pazmino et al. (6), three factors were identified using exploratory factor analysis on nine variables: Patient-Reported (PRF), Clinical (CF), and Laboratory (LF) factor. Factor loadings, which represent how strong a variable relates to its factor, from the exploratory factor analyses (EFAs) were used as weights. The individual variables were normalised to a 0-1 scale considering clinically feasible maximum and minimum. Afterwards, the normalised (n.) variables were weighted-multiplied by the factor loadings-described as follows to calculate:

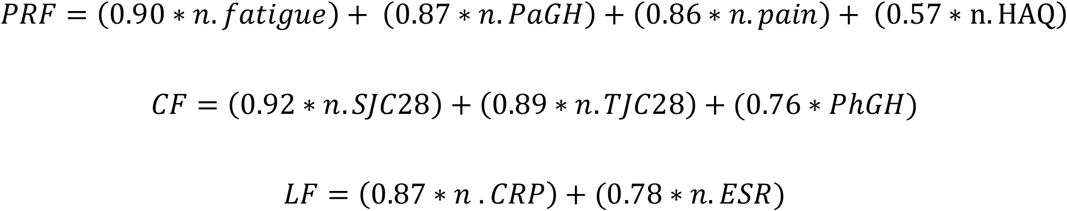

Because the number of variables was different for each factor, the PRF, CF and LF factor scores were re-scaled to 0-1 (higher values suggest more health impact) for comparisons. Thus, for each patient three factor scores per visit were obtained.

Furthermore, a discordance score (DS) for the PRF and CF/LF factor scores was calculated by subtracting the mean of the other two factor scores from the PRF score:

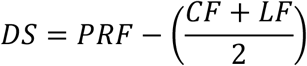

The higher the score, the higher the patient-experienced impact not addressed by traditional measures of disease activity. Correlations between all three factor scores and discordance scores per timepoint were calculated to assess the strength of the relationship of the factor and discordance scores at different time points. Due to the skewed distribution of the factor scores, Spearman correlations were used.

Furthermore, the percentage (%) of improvement from baseline to week 104 and the area under the curve (AUC) across time points were calculated per score. Differences in % improvement and AUC were compared between patients not achieving and achieving early and sustained (week 16 to week 104) disease activity score remission (DAS28CRP <2.6) with ANOVA. Bonferroni correction was used for multiple testing. We chose to look at patients achieving early and sustained remission as a surrogate for “good responders” in whom we expected disease burden to be less pronounced.^12, 15^ To find a clinically meaningful cut-off for the discordance score to serve as an alarm system for unmet patient needs, we based ourselves on the SF-36. Receiver Operator Characteristics (ROC) Curves were fitted for the discordance score to be able to clasify patients into good/poor health status based on SF-36. Without referring to norms, anytime a scale score is below 50, health status is below average.^14^ Logistic regression was fitted to predict sustained remission based on factor and discordance scores at earlier time points with and without adjustment for age and sex. Linear regressions to predict RA-QoL at week 52 and 104 based on the factor scores was also employed.

All analyses were performed with R V.4.1.2.

## RESULTS

Patients with early RA (n=379) were included with a mean (SD) age of 53.9 (13.0), 77% positive to RF or anti-CCP and 69% women. In total, 289 were stratified to high risk and 90 to low risk.

PRF, CF and LF scores improved rapidly over the first 8 weeks (Figure 1a). From baseline to week 104 the scores improved 41%, 78% and 10% for the PRF, CF and LF respectively in the entire population (n=379), 57%, 90% and 27% in patients achieving sustained remission (n=122), and 32%, 77% and 9% in patients not achieving sustained remission (n=257). After correction for multiple testing, there was a statistically significant difference in CF (p<0.001) but not for PRF (p=0.13) and LF (p=0.36) between patients in sustained remission or not (Figure 1b). Figure 1 also indicates higher values of PRF scores at all times, while CF and LF scores are rather similar, which illustrates the rationale for using the discordance score. Further detailed information can be found in Table 1.

**Table 1:**
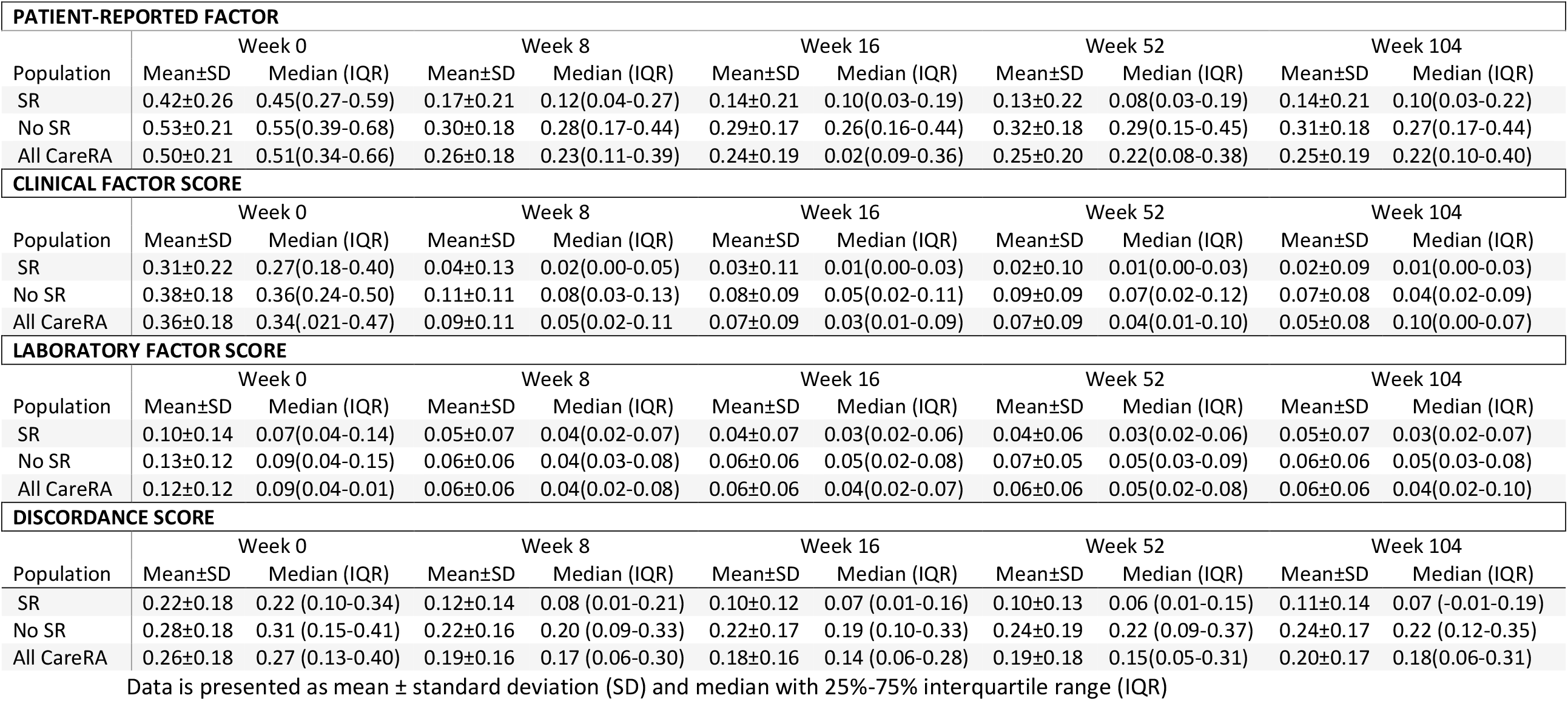
Factors and discordance score per relevant time point in the total population and in patients achieving sustained remission or not (DAS28CRP<2.6 from week16 to104)R

| PATIENT-REPORTED FACTOR |  |  |  |  |  |  |  |  |  |  |
| --- | --- | --- | --- | --- | --- | --- | --- | --- | --- | --- |
|  | Week 0 |  | Week 8 |  | Week 16 |  | Week 52 |  | Week 104 |  |
| Population | Mean±SD | Median (IQR) | Mean±SD | Median (IQR) | Mean±SD | Median (IQR) | Mean±SD | Median (IQR) | Mean±SD | Median (IQR) |
| SR | 0.42±0.26 | 0.45(0.27-0.59) | 0.17±0.21 | 0.12(0.04-0.27) | 0.14±0.21 | 0.10(0.03-0.19) | 0.13±0.22 | 0.08(0.03-0.19) | 0.14±0.21 | 0.10(0.03-0.22) |
| No SR | 0.53±0.21 | 0.55(0.39-0.68) | 0.30±0.18 | 0.28(0.17-0.44) | 0.29±0.17 | 0.26(0.16-0.44) | 0.32±0.18 | 0.29(0.15-0.45) | 0.31±0.18 | 0.27(0.17-0.44) |
| All CareRA | 0.50±0.21 | 0.51(0.34-0.66) | 0.26±0.18 | 0.23(0.11-0.39) | 0.24±0.19 | 0.02(0.09-0.36) | 0.25±0.20 | 0.22(0.08-0.38) | 0.25±0.19 | 0.22(0.10-0.40) |
| CLINICAL FACTOR SCORE |  |  |  |  |  |  |  |  |  |  |
|  | Week 0 |  | Week 8 |  | Week 16 |  | Week 52 |  | Week 104 |  |
| Population | Mean±SD | Median (IQR) | Mean±SD | Median (IQR) | Mean±SD | Median (IQR) | Mean±SD | Median (IQR) | Mean±SD | Median (IQR) |
| SR | 0.31±0.22 | 0.27(0.18-0.40) | 0.04±0.13 | 0.02(0.00-0.05) | 0.03±0.11 | 0.01(0.00-0.03) | 0.02±0.10 | 0.01(0.00-0.03) | 0.02±0.09 | 0.01(0.00-0.03) |
| No SR | 0.38±0.18 | 0.36(0.24-0.50) | 0.11±0.11 | 0.08(0.03-0.13) | 0.08±0.09 | 0.05(0.02-0.11) | 0.09±0.09 | 0.07(0.02-0.12) | 0.07±0.08 | 0.04(0.02-0.09) |
| All CareRA | 0.36±0.18 | 0.34(0.21-0.47) | 0.09±0.11 | 0.05(0.02-0.11) | 0.07±0.09 | 0.03(0.01-0.09) | 0.07±0.09 | 0.04(0.01-0.10) | 0.05±0.08 | 0.10(0.00-0.07) |
| LABORATORY FACTOR SCORE |  |  |  |  |  |  |  |  |  |  |
|  | Week 0 |  | Week 8 |  | Week 16 |  | Week 52 |  | Week 104 |  |
| Population | Mean±SD | Median (IQR) | Mean±SD | Median (IQR) | Mean±SD | Median (IQR) | Mean±SD | Median (IQR) | Mean±SD | Median (IQR) |
| SR | 0.10±0.14 | 0.07(0.04-0.14) | 0.05±0.07 | 0.04(0.02-0.07) | 0.04±0.07 | 0.03(0.02-0.06) | 0.04±0.06 | 0.03(0.02-0.06) | 0.05±0.07 | 0.03(0.02-0.07) |
| No SR | 0.13±0.12 | 0.09(0.04-0.15) | 0.06±0.06 | 0.04(0.03-0.08) | 0.06±0.06 | 0.05(0.02-0.08) | 0.07±0.05 | 0.05(0.03-0.09) | 0.06±0.06 | 0.05(0.03-0.08) |
| All CareRA | 0.12±0.12 | 0.09(0.04-0.01) | 0.06±0.06 | 0.04(0.02-0.08) | 0.06±0.06 | 0.04(0.02-0.07) | 0.06±0.06 | 0.05(0.02-0.08) | 0.06±0.06 | 0.04(0.02-0.10) |
| DISCORDANCE SCORE |  |  |  |  |  |  |  |  |  |  |
|  | Week 0 |  | Week 8 |  | Week 16 |  | Week 52 |  | Week 104 |  |
| Population | Mean±SD | Median (IQR) | Mean±SD | Median (IQR) | Mean±SD | Median (IQR) | Mean±SD | Median (IQR) | Mean±SD | Median (IQR) |
| SR | 0.22±0.18 | 0.22 (0.10-0.34) | 0.12±0.14 | 0.08 (0.01-0.21) | 0.10±0.12 | 0.07 (0.01-0.16) | 0.10±0.13 | 0.06 (0.01-0.15) | 0.11±0.14 | 0.07 (-0.01-0.19) |
| No SR | 0.28±0.18 | 0.31 (0.15-0.41) | 0.22±0.16 | 0.20 (0.09-0.33) | 0.22±0.17 | 0.19 (0.10-0.33) | 0.24±0.19 | 0.22 (0.09-0.37) | 0.24±0.17 | 0.22 (0.12-0.35) |
| All CareRA | 0.26±0.18 | 0.27 (0.13-0.40) | 0.19±0.16 | 0.17 (0.06-0.30) | 0.18±0.16 | 0.14 (0.06-0.28) | 0.19±0.18 | 0.15(0.05-0.31) | 0.20±0.17 | 0.18(0.06-0.31) |

Data is presented as mean ± standard deviation (SD) and median with 25%-75% interquartile range (IQR)

**Figure 1:**
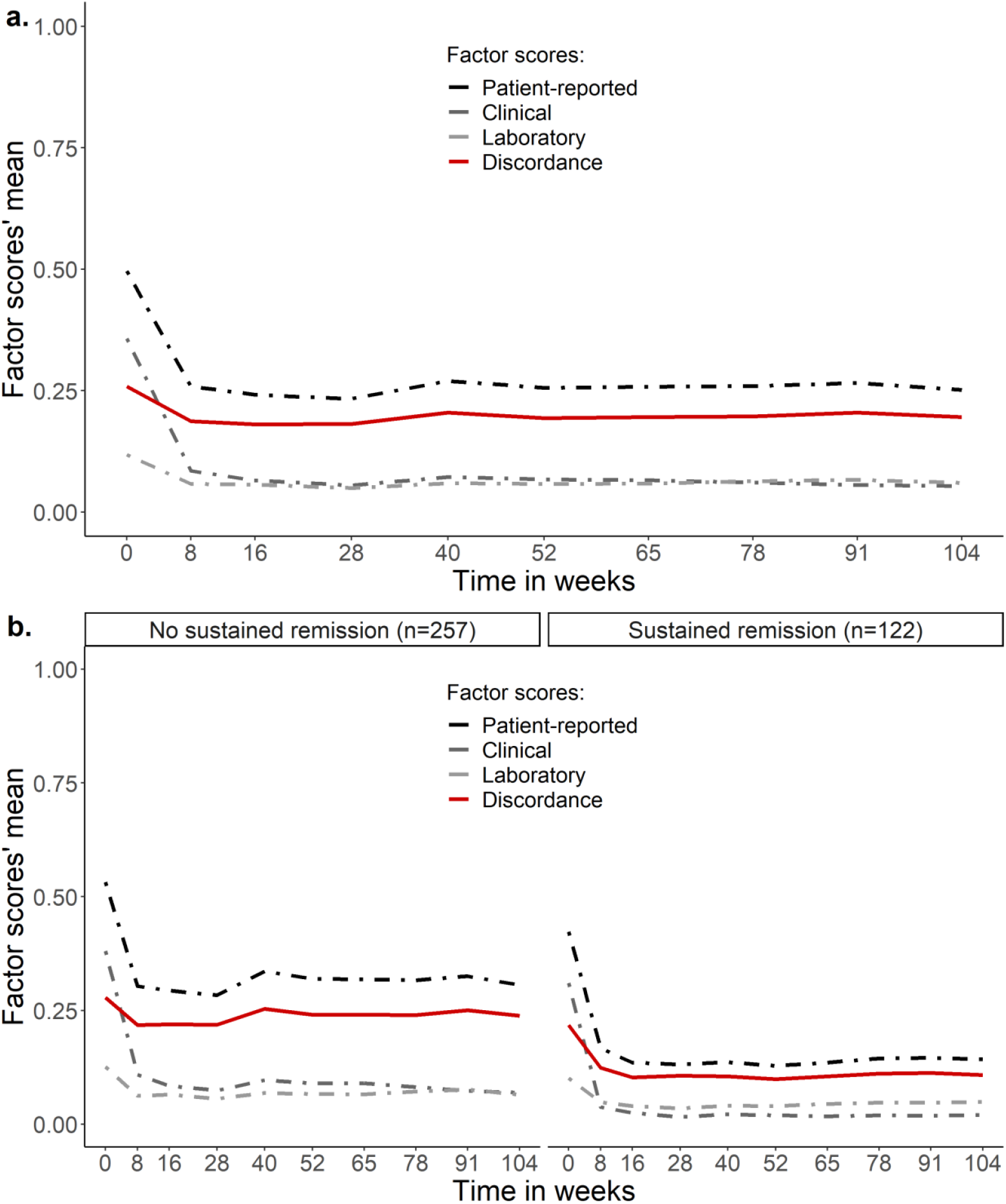
Mean factor score evolution over the 2-year CareRA trial for a. the entire population and b. divided in patients achieving sustained remission or not (DAS28CRP<2.6)

Patients in CareRA had an AUC of 27.4, 7.9 and 6.4 for PRF, CF, LF scores respectively in the overall population. Those who achieved sustained remission had an AUC of 15.6, 3.4 and 4.8 for the PRF, CR and LF scores respectively, compared to 33.2, 10.1, and 7.2 in participants not achieving sustained remission (p<0.001 for all AUCs, between patients in sustained remission or not).

Supplemental Figure 1 shows the Spearman correlations of all three factor scores and their discordance scores at every timepoint. The correlations indicate a strong relationship between the discordance scores at an early stage and both factor scores and the discordance score at a later time point.

Furthermore, PRF was predictive of sustained remission from as early as baseline (Table 2) in univariate and multivariate logistic regression models even after correcting for age and gender. The odds ratio in the multivariate model for PRF was negative meaning that higher values in the PRF score were associated with a lower probability of achieving sustained remission.

**Table 2:**
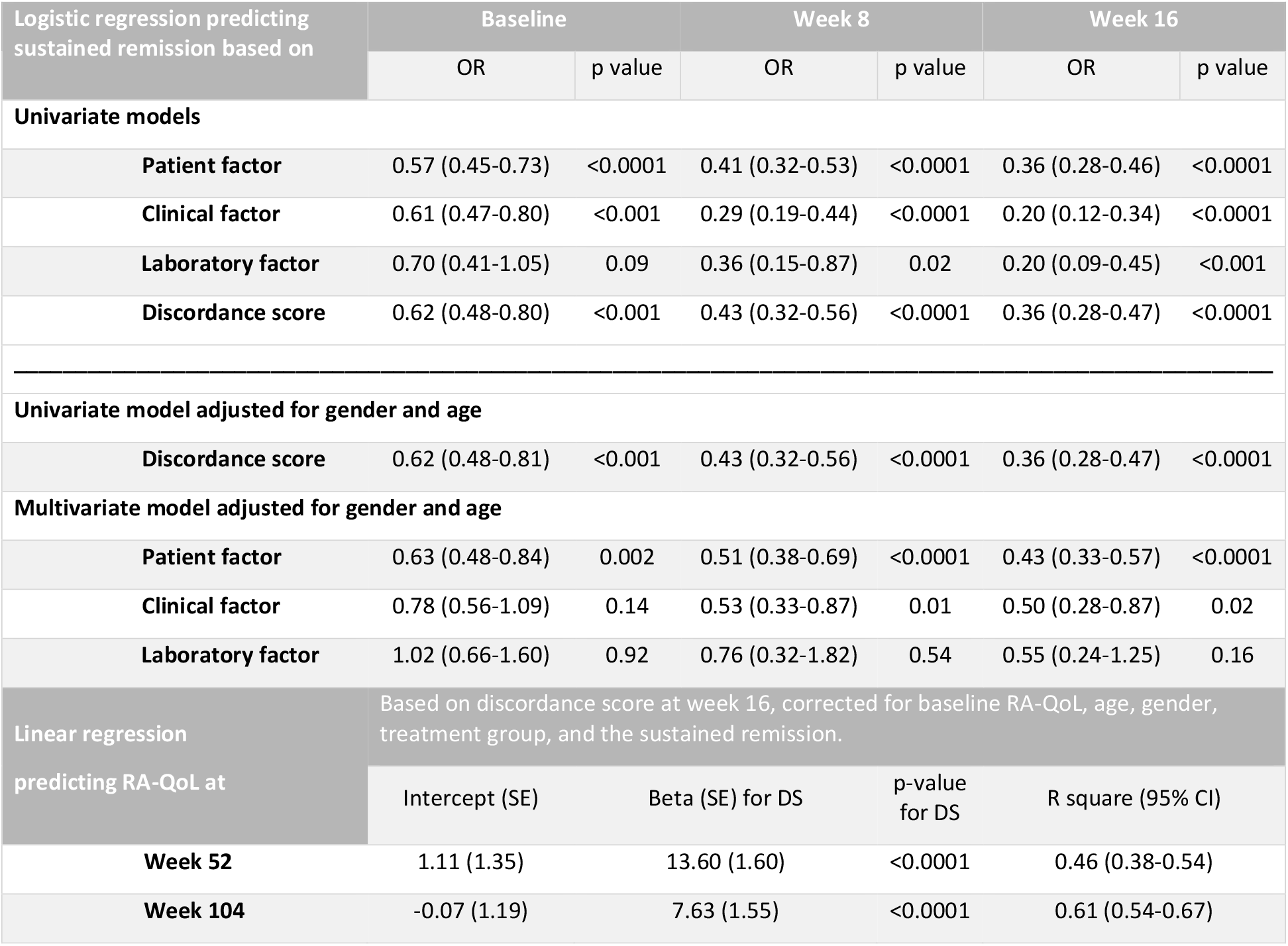
Predictive value of the different factor and discordance scores at baseline, week 8 and 16 for predicting sustained remission (DAS28CRP<2.6 from week 16 to 104) via logistic regression and RA-related quality of life (week 52 and 104) via linear regression.

Additionally, the discordance score proved to be predicting RA-related QoL both at week 52 and week 104 (p < 0.0001) indicating the efficacy of the predictive value of the DS for an unrelated outcome measure.

Regarding the exploratory ROC curve analysis, the area under the curve for every timepoint is around 0.80. Two cut-offs are needed, one for baseline (right before treatment has started) which was estimated at 0.23 and another for when treatment has taken effect (week 16, 52, 104) of around 0.10 to 0.15 (Figure 2).

**Figure 2:**
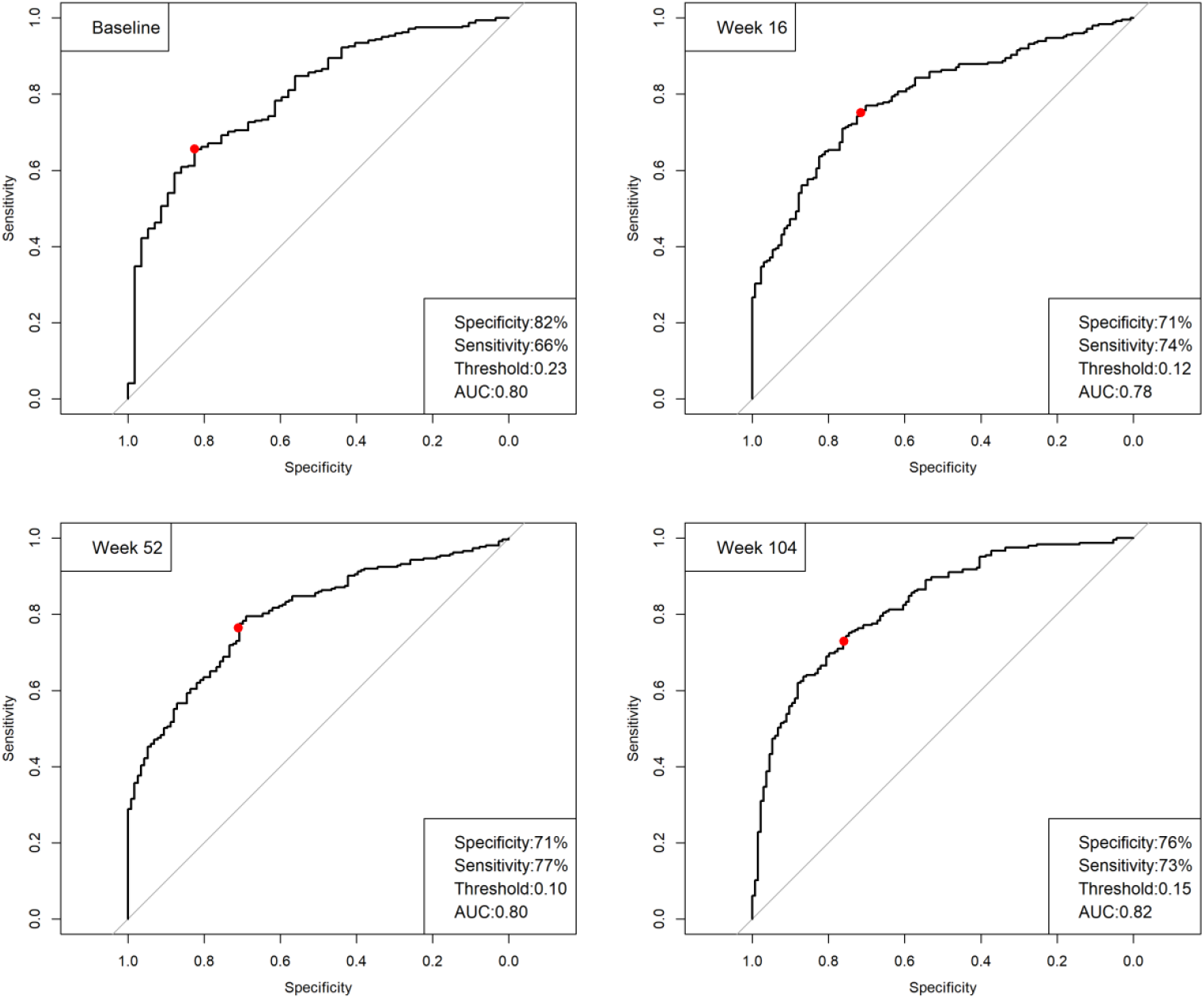
Receiver Operator Characteristics curves for classifying patients based on the discordance score into good/bad health status (based on SF-36) at baseline, week 16, 52, and 104.

## DISCUSSION

The PRF, CF and LF scores improved rapidly over time in a treat-to-target setting in patients with early RA participating in the CareRA trial. However, overall, patient-reported impact seemed not to improve to the same extent. In fact, patient-reported scores remained in all timepoints higher than either CF or LF scores, despite being normalised to the same scale. Furthermore, the difference seems to be constant over time after week 8. This means that assessment of the discordance between PRF scores and the other factor scores may (from already very early in the treatment process) be used as a **warning system** or perhaps even further developed as a **clinical decision support tool** for the clinician since it predicts future patient-reported impact. Even more importantly, it could detect at an earlier stage unmet needs even in patients “under control”, considering that one in five persistent treatment responders from CareRA reported unmet needs after 1 year, such as remaining pain and fatigue.^13^ The fact that two cut-offs are needed for the discordance score seems to indicate that there is a different way of evaluationg patient’s unmet needs after recently being diagnosed (baseline) versus when treatment has been started. More validation is needed.

Moreover, the PRF score also has a predictive value for forecasting if a patient will be achieving sustained remission. These findings may be somewhat limited since the analyses were exploratory and not powered for assessing prediction models based on the factor and discordance scores. Hence, there could be an effect of the LF which could not be captured due to a too low sample size. The probability of achieving a state of disease control decreases from baseline onwards for every point increase in the PRF score and the negative predictive value of the PRF score becomes even more important by week 16, highlighting again the importance of early disease control. Even perhaps early control of the global disease impact, which most likely is not achieved with medication only. Keeping an open mind to the alternative that complementary interventions beside DMARDs could help mitigate the “persisting effect of disease” is crucial. This is an important issue for future research. Studies in other patient cohorts than the one the scores were developed in, are needed for future validation of these factor scores, as well as qualitative studies to understand what is behind this discordance between patient reported outcomes and clinical variables, and possible cut-offs of the discordance score for further clinical use.

Rapid and persistent disease control as well as baseline psychosocial variables, and not so much treatment choice, have been associated with favourable patient-reported health and illness perceptions after 1 year in CareRA. ^13^ The early, intensive, treat-to-target approach^3^ definitely has its benefits with snowballing consequences that can go further than controlling disease activity.^15^ However, there might be space for broadening the target currently used.^3^

## CONCLUSION

Patients’ unmet needs in terms of pain, fatigue, functionality and overall well-being should be given more attention during follow-up, even in individuals achieving sustained remission. Looking at the discordance between the Patient factor score and the Clinical and Laboratory scores does provide further insights in these needs, allowing to broaden the future scope of the treat-to-target principle to multidisciplinary interventions on top of, and sometimes as an alternative for pharmacological treatment adaptations.

## Data Availability

The authors commit to making the relevant anonymised patient data available for a specified purpose approved by the institution and the principal investigator of the CareRA study and with a signed data access agreement.

---

**Supplemental Figure 1:**
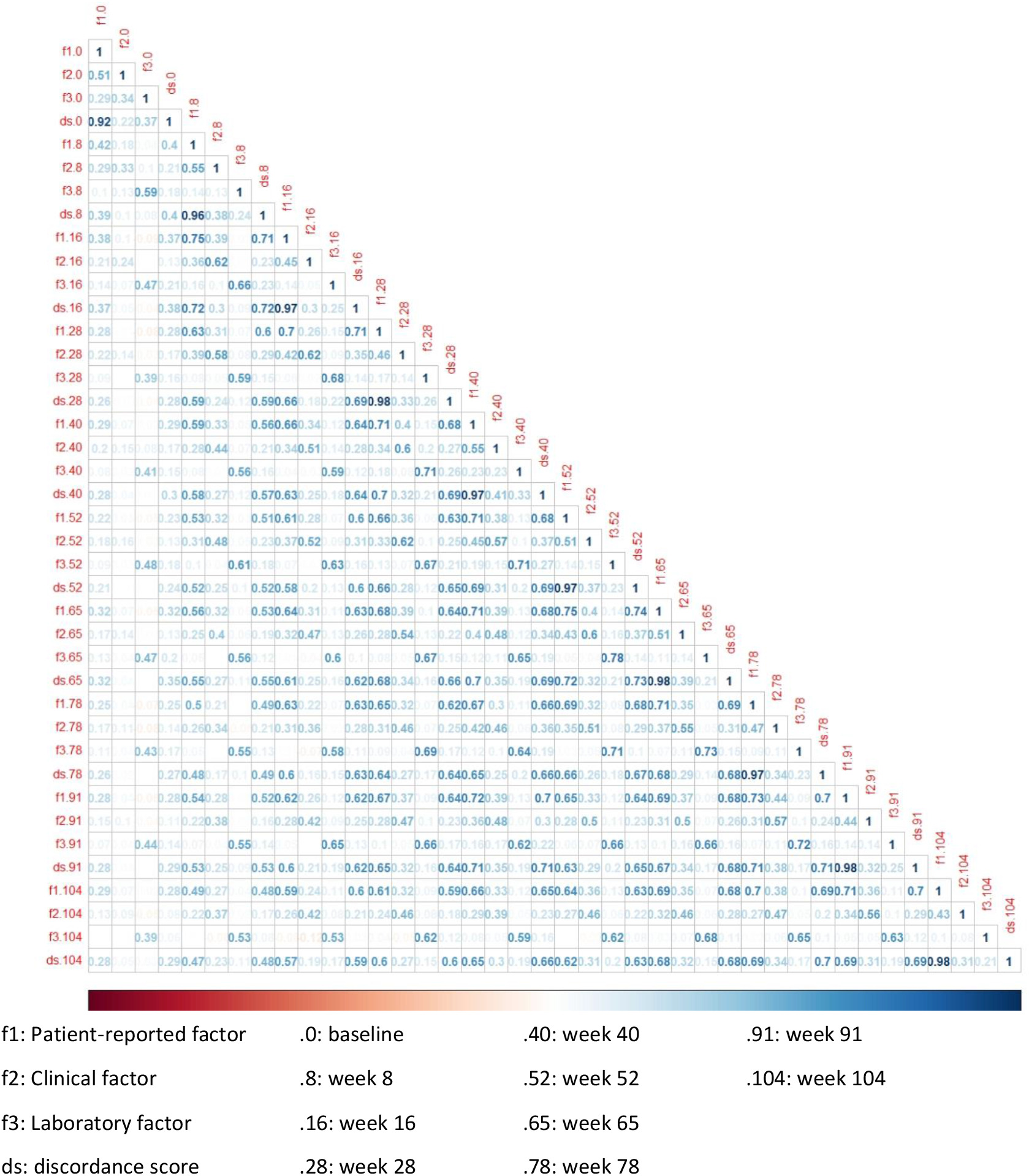
Spearman correlations of the factor scores and discordance scores at every timepoint The color scheme for the correlations goes from −1 (red) to +1 (blue). This is a colored representation of the strength of each correlation.

## Acknowledgements

We would like to show our gratitude to all participating patients, as well as to the investigators and medical staff at all sites. We appreciate the time invested.

## Contributors

PV, RW, AB, SP, and AL made substantial contributions to the conception or design of the study. SP and AL performed the statistical analysis. The manuscript was written by SP, AL, PV, RW, and AB and subsequently revised critically by all the remaining co-authors. All authors were involved in data interpretation and approved the final version to be submitted for publication.

## Patient consent for publication

Not required.

## Ethics Approval

The study was approved by the leading Ethics Committee of the University Hospitals Leuven after consulting the medical ethics committee of each participating centre (ref s51411), and all study participants gave their written informed consent before inclusion.

## Provenance and peer review

Not commissioned; externally peer-reviewed.

## Patient involvement

The pragmatic CareRA protocol was strongly inspired by daily interactions of the investigators with RA patients in daily clinical practice. Patients were not formally involved in setting the research question or the outcome measures, nor were they invited to comment on study design or the interpretation of the results of this manuscript. However, results of this research will be disseminated to study participants, all stakeholders and the general public in collaboration with patient organisations and the Belgian patient partners program (trained patients who educate physicians, medical students and other health care professionals in collaboration with a rheumatologist).

## REFERENCES

1. Scott, D. L., Wolfe, F. & Huizinga, T. W. J. Rheumatoid arthritis. in The Lancet vol. 376, 1094–1108 (2010).

2. Van Riel, P. L. C. M. The development of the disease activity score (DAS) and the disease activity score using 28 joint counts (DAS28). Clin Exp Rheumatol vol. 32 http://www.clinexprheumatol.org/article.asp?a=8657 (2014).

3. Smolen, J. S. et al. EULAR recommendations for the management of rheumatoid arthritis with synthetic and biological disease-modifying antirheumatic drugs: 2019 update. Ann. Rheum. Dis. 79, 685–699 (2020).

4. Smolen, J. S. et al. Treating rheumatoid arthritis to target: Recommendations of an international task force. Ann. Rheum. Dis. 69, 631–637 (2010).

5. Ferreira, R. J., Landewé, R.B. & da Silva, J. A. Definition of Treatment Targets in Rheumatoid Arthritis: Is It Time for Reappraisal? J. Rheumatol. (2021) doi:10.3899/jrheum.210050.

6. Ferreira, R. J. de O. et al. Drivers of patient global assessment in patients with rheumatoid arthritis who are close to remission: An analysis of 1588 patients. Rheumatol. (United Kingdom) 56, 1573–1578 (2017).

7. Ferreira, R. J. O. et al. Impact of Patient’s Global Assessment on Achieving Remission in Patients With Rheumatoid Arthritis: A Multinational Study Using the METEOR Database. Arthritis Care Res. 71, 1317–1325 (2019).

8. Ferreira, R. J. O. et al. Suppressing Inflammation in Rheumatoid Arthritis: Does Patient Global Assessment Blur the Target? A Practice-Based Call for a Paradigm Change. Arthritis Care Res. 70, 369–378 (2018).

9. Radner, H. et al. Different Rating of Global Rheumatoid Arthritis Disease Activity in Rheumatoid Arthritis Patients With Multiple Morbidities. Arthritis Rheumatol. 69, 720–727 (2017).

10. Pazmino, S. et al. Does Including Pain, Fatigue, and Physical Function When Assessing Patients with Early Rheumatoid Arthritis Provide a Comprehensive Picture of Disease Burden? J. Rheumatol. Nov, jrheum.200758 (2020).

11. Pazmino, S. et al. Evaluation of disease burden by (separate) patient-reported, clinical and laboratory factor-scores in patients with established rheumatoid arthritis: factor analysis replication J. Rheumatol, in press.

12. Stouten, V. et al. Effectiveness of different combinations of DMARDs and glucocorticoid bridging in early rheumatoid arthritis: two-year results of CareRA. Rheumatology (Oxford). (2019) doi:10.1093/rheumatology/kez213.

13. Van Der Elst, K. et al. One in five patients with rapidly and persistently controlled early rheumatoid arthritis report poor well-being after 1 year of treatment. RMD Open 6,(2020).

14. Van der Elst, K. et al. Patient reported outcome data from the Care in Early Rheumatoid Arthritis trial: Opportunities for broadening the scope of treating to target. Arthritis Care Res. (Hoboken). (2019) doi:10.1002/acr.23900.

15. Schoemaker, C. G. & de Wit, M. P. Treat to target from the patient perspective is bowling for a perfect strike. Arthritis Rheumatol. part.41461 (2020) doi:10.1002/art.41461.

